# The spatio-temporal distribution of COVID-19 infection in England between January and June 2020

**DOI:** 10.1101/2021.02.22.21251534

**Authors:** Richard Elson, Tilman M. Davies, Iain R. Lake, Roberto Vivancos, Paula B. Blomquist, Andre Charlett, Gavin Dabrera

## Abstract

The spatio-temporal dynamics of an outbreak provide important insights to help direct public health resources intended to control transmission. They also provide a focus for detailed epidemiological studies and allow the timing and impact of interventions to be assessed.

A common approach is to aggregate case data to administrative regions. Whilst providing a good visual impression of change over space, this method masks spatial variation and assumes that disease risk is constant across space. Risk factors for COVID-19 (e.g. population density, deprivation and ethnicity) vary from place to place across England so it follows that risk will also vary spatially. Kernel density estimation compares the spatial distribution of cases relative to the underlying population, unfettered by arbitrary geographical boundaries, to produce a continuous estimate of spatially varying risk.

Using test results from healthcare settings in England (Pillar 1 of the UK Government testing strategy) and freely available methods and software, we estimated the spatial and spatio-temporal risk of COVID-19 infection across England for the first six months of 2020. Widespread transmission was underway when partial lockdown measures were introduced on the 23^rd^ March 2020 and the greatest risk erred towards large urban areas. The rapid growth phase of the outbreak coincided with multiple introductions to England from the European mainland. The spatio-temporal risk was highly labile throughout.

In terms of controlling transmission, the most important practical application is the accurate identification of areas *within* regions that may require tailored intervention strategies. We recommend that this approach is absorbed into routine surveillance outputs in England. Further risk characterisation using widespread community testing (Pillar 2) data is needed as is the increased use of predictive spatial models at fine spatial scales.

## Background

On the 31 December 2019, the World Health Organization (WHO) was informed of a cluster of cases of pneumonia of unknown cause detected in Wuhan City, Hubei Province, China. Since the initial identification of SARS-CoV-2 as the cause of COVID-19, over 32 million cases have been diagnosed globally, with more than 900,000 fatalities, as of 27^th^ September 2020 [1]. The first laboratory confirmed case in England was reported on the 31^st^ January 2020. A series of interventions designed to slow rates of infection followed, culminating in a partial lockdown announced by the UK Government on the 23rd March 2020.

Understanding the spatiotemporal dynamics of COVID-19 helps to clarify the extent and impact of the pandemic and can aid decision making, planning and community action intended to control transmission [2]. It also provides an opportunity to assess the impact of interventions over space and time.

One approach is to describe changes in infection rates within administrative boundaries in England have been published widely. This approach expresses the disease risk per head of population and assumes that risk is constant across space i.e. the risk of disease does not depend upon spatial location. This is rarely the case and the distribution of risk factors for COVID-19 (for example population density, deprivation and ethnicity) are known to vary across England so it follows that absolute risk will also vary spatially.

Another approach is to plot points to produce a spatial point pattern. This is useful for small data sets but as the number of points increases, over plotting makes it difficult to discriminate between the relative densities of points.

Kernel density estimation (KDE), also known as kernel smoothing, is a flexible, non-parametric method by which spatially varying risk may be estimated without the need to aggregate data. Smoothing a spatial point pattern (using an appropriate bandwidth) overcomes the over plotting problem by expressing the number of points as an intensity function. Comparing the intensities of two groups, for example those with an infectious disease and those without, across a defined geographical area results in an intensity (or risk) ratio. If the ratio is ∼1, this suggests that the risk of infection is unrelated to spatial location. Evidence of spatial variation in risk occurs where the intensities differ. Ratio values >1 indicate an increased risk and values <1 indicate lower risk.

As the COVID-19 outbreak progresses in England, KDE provides a scalable means to identify areas of significantly higher or lower risk to inform national policy and local action.

Using established methods [3-5] and freely accessible software [3, 6], we conducted a spatio-temporal point pattern analysis of COVID-19 risk in England between January and June 2020. Our aims were to describe the spatio-temporal dynamics of the first six months of the COVID-19 outbreak and assess the potential use of this method to inform and support public health policy decisions as the outbreak progresses.

## Methods

The method we followed is described in detail by Davies et al. [3] and Elson et al. [7]. For the spatial estimates, each set of points (case and control) were smoothed using an adaptive [4, 8] bandwidth to determine the spread of smoothing kernels centred on each point to produce a density surface. Adaptive bandwidths account for greater uncertainty in areas with fewer points (e.g. rural areas) so the bandwidth is large resulting in greater smoothing. In urban areas, more data points mean the bandwidth is smaller resulting in a surface with less smoothing. Calculating the ratio of case and control densities provides a continuous estimate of relative risk which can be plotted on a map [4, 9, 10].

### Case locations

We selected confirmed cases of COVID-19 reported to the PHE Second Generation Surveillance System (SGSS) under Pillar 1 of the UK Government testing strategy between the 31^st^ January 2020 and the 30th June 2020. Pillar 1 includes tests only for those with a medical need (symptomatic and seen by a clinician) but may also include some healthcare workers and samples taken as part of outbreak investigations [11]. The data was checked for duplicates and presence of a valid residential postcode. Postcodes are like US Zip codes and represent a single residential street or group of houses.

The statistical methodology for spatial point processes are very sensitive to duplicate data points [12]. We used unique control locations but included multiple cases with the same postcode to account for sporadic and outbreak cases.

### Population at risk (‘control’) locations

The underlying population at risk (‘controls’) was represented by points randomly sampled from the National Population Database (NPD). The NPD is a Geographical Information System (GIS) dataset that combines multiple layers of data (including population) in a 100-metre by 100-metre grid [13, 14]. Based on the centroid coordinates of each grid square, ‘control’ locations were randomly drawn without replacement. The probability of a location being drawn was weighted by the summed population of each grid square to reflect the spatially varying nature of the underlying population at risk. The number of controls was chosen to match the number of cases.

For the spatial estimates, we attempted four bandwidth initialisation methods to set the ‘global’ and ‘pilot’ smoothing parameters needed to calculate the adaptive bandwidths themselves: maximal smoothing [15], bootstrapping [3, 16], least-squares cross validation (LSCV) [17] and likelihood cross-validation [18, 19].

The resulting bandwidths were used to produce density estimates at all locations of a fine grid of co-ordinates laid within a simplified polygon of the mainland boundary of England and the Isle of Wight.

To explore the temporal variation in the spatial risk, we marked each case with the date that their specimen was taken. For cases with multiple test results, the specimen date that gave the most recent positive result was used. We then calculated the number of days that had elapsed from the specimen date of the first confirmed case (31^st^ January 2020) as the temporal event. The spatiotemporal relative risk surface was then calculated using the fixed estimator of Fernando & Hazelton [20].

All estimates are edge-corrected to account for kernel weight lost over the boundary of the study region [21, 22]and, for the spatial analyses only, are calculated as symmetric adaptive risk function estimates using the pooled case/control data and equal global and pilot bandwidths [5]. Unless stated otherwise, results are reported as log-relative risk surfaces. Contours identifying areas of significantly higher risk were superimposed at the 1% significance level for the spatial estimates and 1% and 0.01% levels for the spatio-temporal estimates. Wherever temporal results are referred to in terms of weeks, this refers to the corresponding International Organization for Standardization (ISO) week.

All analyses were performed using the contributed packages sparr [3] and spatstat [12, 23] in the R language [6].

## Results

Between the 31st January and the 30^th^ June 2020, 160,976 cases of COVID-19 were reported to PHE under Pillar 1 of the UK Government testing strategy [11]. Of these, residential postcodes were available for 154,210 (96%). Of these, multiple cases were recorded at 44,989 (30%) postcode locations.

### Bandwidth selection

The oversmoothed and bootstrap methods produced usable spatial and space-time bandwidths. The LSCV approach did not provide a result and the likelihood-based approach produced a very small bandwidth that resulted in an under smoothed, ‘spiky’ surface.

### Spatial risk

Figure 1 shows the areas in England classified as urban by the Office for National Statistics (ONS) [24]. The relative risk across England during the study period is presented in Figure 2. With some exceptions, the areas with the highest risk tended to be large urban areas.

**Figure 1.**
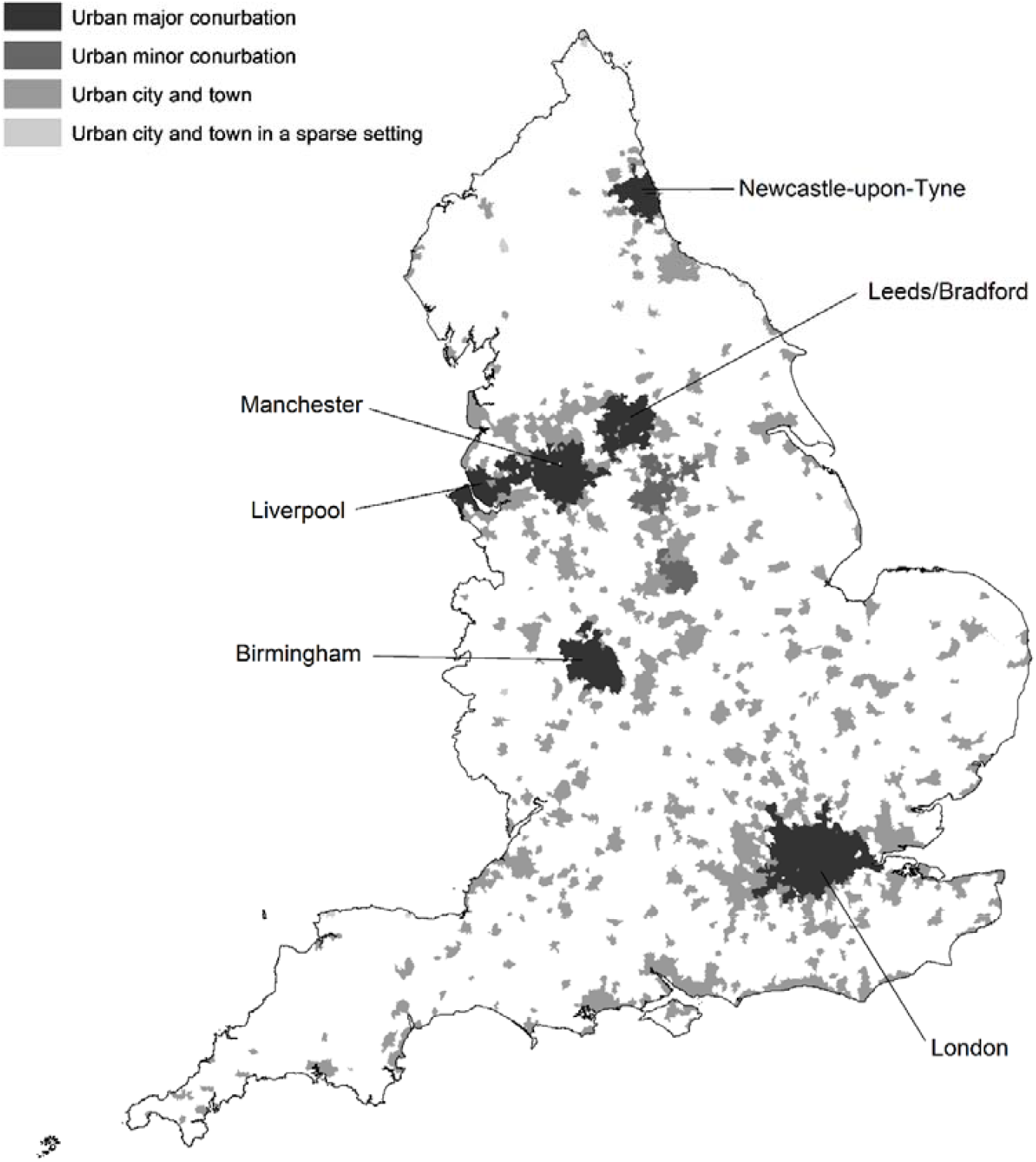
Urban areas in England.

**Figure 2.**
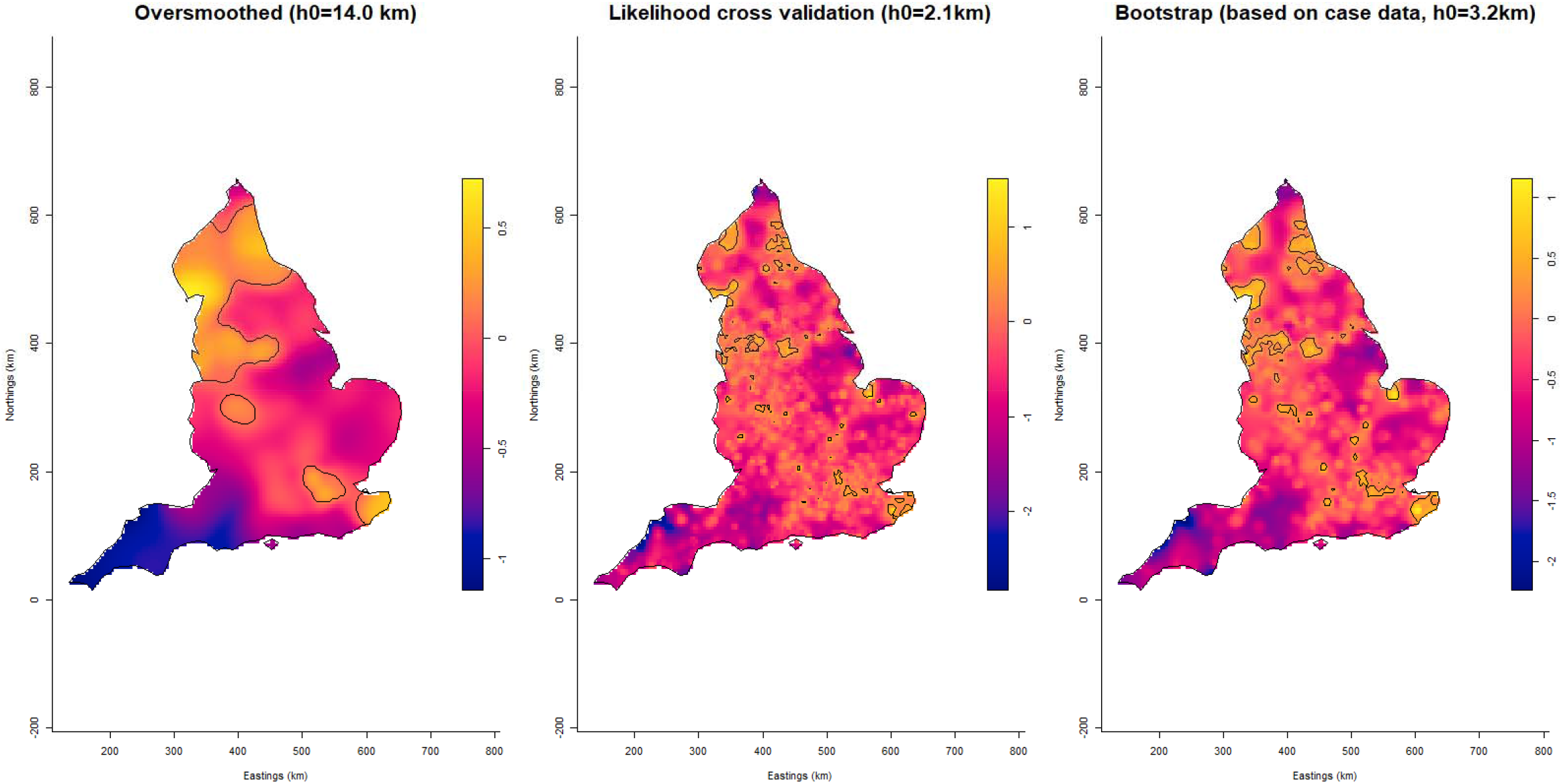
Log relative risk estimates for COVID-19 in England between January and June 2020 using different bandwidths: oversmoothed (left), likelihood cross validation (centre) and bootstrapping (right). Tolerance contours indicating areas of significantly higher risk are superimposed as solid lines at the 1% confidence level

### Spatio-temporal risk

An animation of the spatio-temporal analysis combining an epidemic curve with the risk surfaces using the oversmoothed and bootstrap estimators can be viewed <u>here</u>. The individual risk surface for the oversmoothed version is <u>here</u> and the bootstrap version is <u>here</u>. The 14-day space-time slices are presented in Figures 3 and 4 for the oversmoothed and bootstrap estimators respectively.

**Figure 3.**
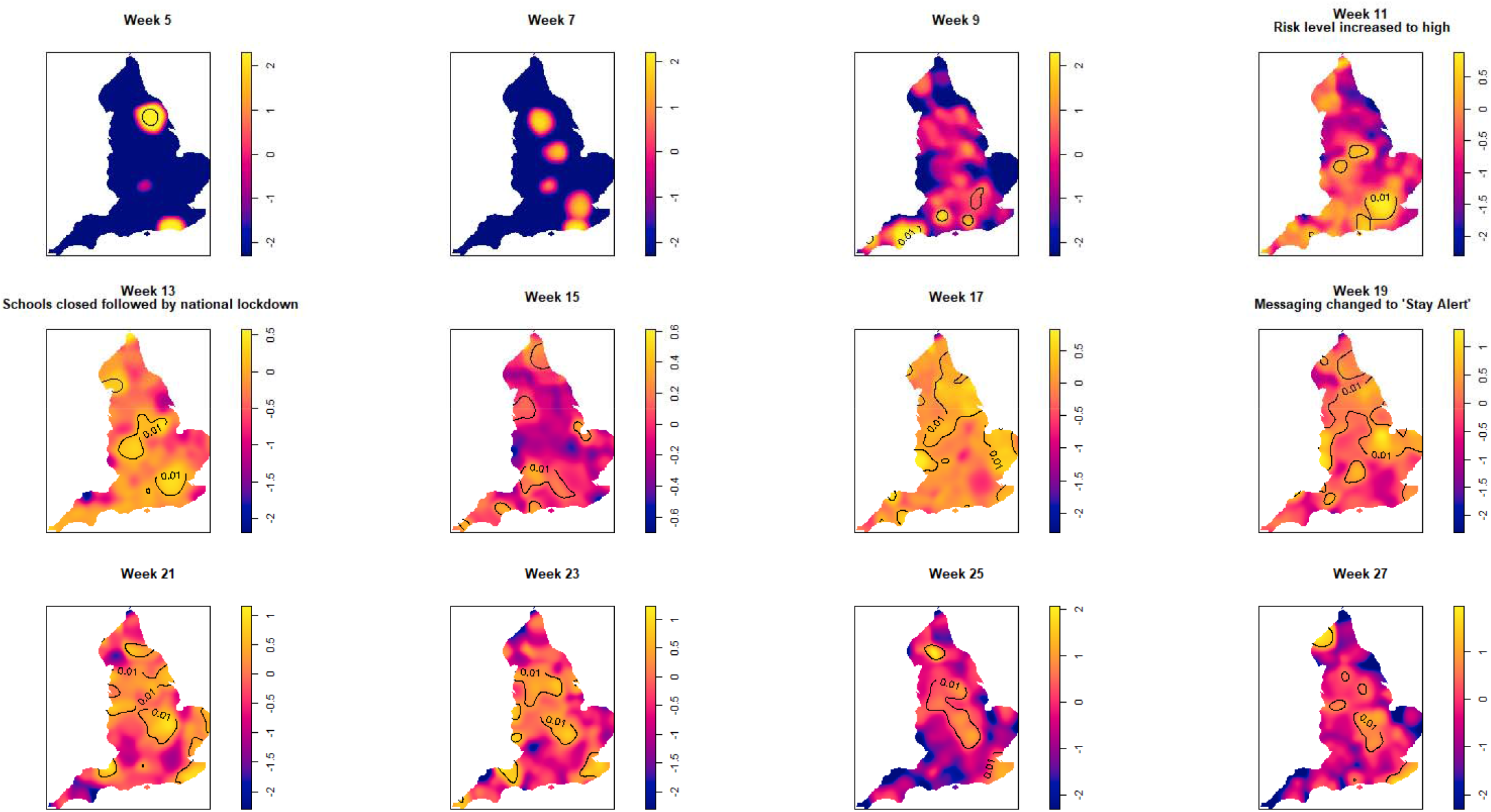
Log relative risk spacetime slices using an oversmoothed bandwidth (*h*=15.4km, *lambda (*λ)=2.04) in 14-day periods from the date of the first case confirmed in ISO Week 5. Solid lines outline areas of significantly higher risk at the 1% confidence level.

**Figure 4.**
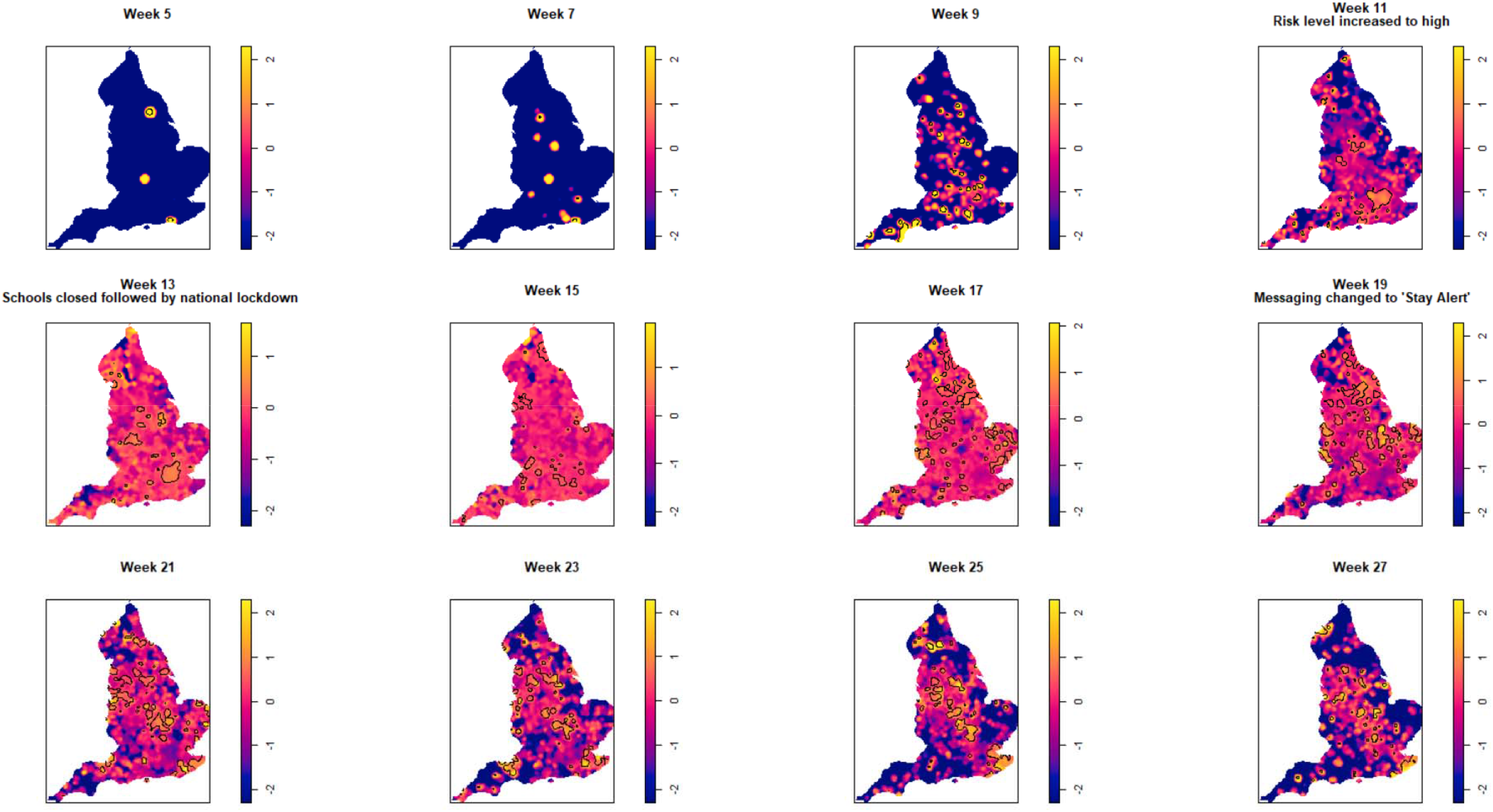
Log relative risk spacetime slices using bootstrap bandwidth (based on cases only: *h*=4.8km, *lambda (*λ) =3.2) in 14-day periods from the date of the first case confirmed in ISO Week 5. Solid lines outline areas of significantly higher risk at the 1% confidence level.

Fewer than twenty cases were recorded between the confirmation of the first case (Week 5 commencing January 27th) and the end of Week 8 (February 23^rd^). Weeks 9 −10 were characterised by a greater geographical spread of small areas of elevated risk and an increase in case numbers to ∼400 by the end of Week 10 (March 8^th^). Week 11 (commencing 9^th^ March) saw a rapid increase in case numbers and significantly elevated risk, particularly in the cities of London and Birmingham. By the end of Week 14 (5th April), 47,668 cases had been reported. From Week 15 (commencing 6^th^ April) onwards, areas of significantly elevated risk became more dispersed with some areas in the North and far South East of the country experiencing sustained periods of elevated risk, even as case numbers declined towards the end of the study period.

Of note is the generalised increase in risk across the country between Weeks 13-19 and the abrupt change in risk seen in London between Weeks 13 and 15.

## Discussion

To the best of our knowledge, this is the first description of the spatio-temporal distribution of COVID-19 in England using unaggregated data. As such, it defines areas of statistically significant high and low risk at a very fine spatial scale, unhampered by administrative boundaries.

Taking into account a seven-day lag for the incubation period [25] prior to sample collection, our results show that geographically widespread transmission was underway at least one week prior to the partial lockdown announced on the 23rd March 2020.

The rapid increase in cases and geographical spread in risk coincided with the roll out of the roll out of PCR assays to hospitals during March resulting in greater ascertainment. However, intensive sequencing of SARS-CoV-2 genomes revealed that there were multiple introductions from European countries. The frequency of these imports (introduced via multiple entry points by travellers returning to the UK predominately from Spain, Italy and France) reached a peak in mid-March 2020 (Week 12) and led to widespread onward transmission within the UK [26].

The risk was greatest in some, but not all, large urban areas. At the beginning of the outbreak, the risk in London was significantly elevated for a prolonged period but changed abruptly within the period of a single week (Week 15). The reasons for this are unclear but may be related to the impact of non-pharmaceutical interventions (social-distancing, reduced use of public transport etc.) or factors related to immunity. Seroprevalence of antibodies to SARS-CoV-2 in samples from healthy adult blood donors in England showed that the prevalence in London, adjusted for assay accuracy, age and sex, increased from 1.5% in Week 13 to 12.3% in Weeks 15 to 16 and 17.5% in Week 18. Given that the antibody response takes at least two weeks to become detectable, those displaying a positive result in Week 18 are likely to have become infected before mid-April. By the end of our study period (Week 27), prevalence had dropped to 10% in London [27].

Large urban areas in England have higher population densities and tend to have higher numbers of black, Asian and minority ethnic residents. They are also the areas with the highest deprivation and air pollution scores: all factors associated with an increased risk of infection and/or poorer outcomes following infection with SARS-CoV 2 [28-30].

Selecting the ‘right’ bandwidth is crucial for this approach. Calculation of the over smoothing bandwidth is extremely quick, and the results provide a good overview of elevated risk.

However, this somewhat rudimentary approach is unlikely to identify focused hotspots. The bootstrap method, whilst more computationally intensive, produced a usable bandwidth in less than thirty minutes for the spatial analysis and around ten hours for the spatio-temporal bandwidth. The resulting output provides superior geographical detail allowing resources to be targeted more efficiently. Notwithstanding this, too small a bandwidth results in an under smoothed surface which can erroneously identify ‘significant’ peaks in risk as a result of increased variability of the kernel estimator. The numeric stability of LSCV and likelihood based methods is known to be questionable in practice, with resulting estimates often being under-smoothed[9].

There are some limitations to this analysis. First, our approach was exploratory and does not account for groups more likely to experience poorer outcomes following infection due to socio-demographic, occupational and environmental factors. Also, the data we used represents those who were symptomatic and sought healthcare. In common with all surveillance systems, this is biased towards the severe end of the disease spectrum. One way of overcoming this bias would be to include results from the wider community testing performed under Pillar 2 of the UK testing regime. We decided not to include this data because Pillar 2 testing was introduced part way through the study and was also subject to data quality issues until May 2020 [11]. The decline in cases described here is likely to be an underestimate of the true community incidence and may not reflect the spatial locations of cases identified under Pillar 2. This requires further investigation, however, considering the way that SARS-CoV-2 is transmitted, we anticipate that this will not differ considerably, and our analysis represents the spatio-temporal ‘tip of the iceberg’ for COVID-19 in England during the study period. Finally, the PCR assay used by hospitals was rolled out nationally during March 2020 resulting in greatly improved case ascertainment. This coincided with the rapid increase seen during March 2020 so may be an artefact of improved surveillance. However, this does not explain the spatial variation noted beyond the end of March (when the assay was in widespread use) nor the sudden decline in cases in Birmingham and London. Our analysis demonstrates how KDE can identify areas of England where the risk of Covid-19 infection differs significantly. In terms of controlling transmission, the most important practical application is the accurate identification of areas *within* regions that require improved public health messaging or tailored intervention strategies. Spatial modelling can be used to the predict the spread of infection [31, 32] and the methodology to do this has been available for some time [32, 33]. Such approaches have already been applied to COVID-19 case data [34] and self-reported symptoms [33]. It’s hoped that this will form part of the UK response strategy in the coming months and will be most informative at very fine spatial scales [33]. To harness the benefits of such modelling approaches, public health organisations and academic centres must find ways to share information and promote collaboration without compromising patient confidentiality.

To conclude, we present a spatio-temporal analysis of COVID-19 in England covering the first six months of 2020. We recommend that this approach is absorbed into routine surveillance outputs and that ways to confidentially share patient data with academic collaborators are explored. Further work using Pillar 2 test data and the development of predictive spatial models at fine spatial scales is needed.

## Supporting information

Spatio-temporal animation 1

Spatio-temporal animation 2

Spatio-temporal animation 3

## Data Availability

The data used to perform this analysis contains Patient Confidential Data information in the
form of geographical coordinates describing a cases place of residence and has not been
made available. PHE has delegated authority, on behalf of the Secretary of State, to process
Patient Confidential Data under Regulation 3 The Health Service (Control of Patient
Information) Regulations 2002
http://www.legislation.gov.uk/uksi/2002/1438/regulation/3/made.
Regulation 3 makes provision for the processing of patient information for the recognition,
control and prevention of communicable disease and other risks to public health.

## Acknowledgements

Thanks are due to all Public Health England staff and the many other professionals who continue to work on the COVID-19 epidemic in England and around the world. Particular thanks are extended to Kate Twohig and Asad Zaidi who provided the data to conduct this study. We are also indebted to two anonymous reviewers for their comments which resulted in a much-improved final manuscript. Finally, thanks are due to the R community who share their knowledge, skills and intellect freely.

RE and RV are affiliated to the National Institute for Health Research Health Protection Research Unit (NIHR HPRU) in Gastrointestinal Infections.

## Ethics declaration

The PHE Research Support and Governance Office exempted this study from full ethical review. The research was classified as surveillance undertaken as part of PHE’s legal responsibility to monitor COVID-19 and was found to be fully compliant with all current regulatory requirements.

## Data availability statement

The data used to perform this analysis contains Patient Confidential Data information in the form of geographical coordinates describing a cases’ place of residence and has not been made available. PHE has delegated authority, on behalf of the Secretary of State, to process Patient Confidential Data under Regulation 3 The Health Service (Control of Patient Information) Regulations 2002 http://www.legislation.gov.uk/uksi/2002/1438/regulation/3/made.

Regulation 3 makes provision for the processing of patient information for the recognition, control and prevention of communicable disease and other risks to public health.

